# Pelvic Pain: what are the symptoms and predictors for surgery, endometriosis and endometriosis severity?

**DOI:** 10.1101/2021.01.29.21250806

**Authors:** Isabelle Conroy, Samantha S Mooney, Shane Kavanagh, Michael Duff, Ilona Jakab, Katharine Robertson, Amy L Fitzgerald, Alexandra Mccutchan, Siana Madden, Sarah Maxwell, Shweta Nair, Nimita Origanti, Alish Quinless, Kelly Mirowska-Allen, Megan Sewell, Sonia R Grover

## Abstract

**Background:** Chronic pelvic pain (CPP) is a common condition which significantly impacts the quality of life and wellbeing of many women.

Laparoscopy with histopathology is recommended for investigation of pelvic pain and identification of endometriosis with concurrent removal. Never-the-less, the association between endometriosis and pelvic pain is challenging, with endometriosis identified in only 30-50% of women with pain.

**Aims:** To explore the predictors for undergoing surgery, for identifying endometriosis and endometriosis severity in a cohort of women with CPP.

**Materials and Methods:** This study forms part of the Persistent Pelvic Pain project, a prospective observational cohort study (ANZCTR:ACTRN12616000150448). Women referred to a public gynaecology clinic with pain were randomised to one of 2 gynaecology units for routine care and followed for 36-months with 6-monthly surveys assessing demographics, medical history, quality of life, and pain symptoms measured on a Likert scale. Operative notes were reviewed, and endometriosis staged.

**Results:** Of 471 women recruited, 102 women underwent laparoscopy or laparotomy, of whom 52 had endometriosis (n=37 stage I-II; n=15 stage III-IV). Gynaecology unit, pelvic pain intensity and lower parity were all predictors of surgery (Odds ratio (OR) 0.342; 95%CI 0.209-0.561; OR 1.303; 95%CI: 1.079-1.573; OR 0.767; 95%CI: 0.620-0.949 respectively). There were no predictors identified for endometriosis diagnosis and the only predictor of severity was increasing age (OR 1.155; 95%CI: 1.047-1.310).

**Conclusions:** Pain intensity and gynaecology unit were key predictors of undergoing laparoscopy, however, pain severity did not predict endometriosis diagnosis or staging. These findings indicate the need to review current frameworks guiding practice towards surgery for pelvic pain.

## Introduction

Chronic pelvic pain (CPP) and endometriosis are often seen together in the literature and clinical medicine, but their exact relationship remains unclear. CPP is a common condition, impacting significantly on women, their health, general quality of life and social engagement^1^. Exact definitions differ creating inconsistency in research^2^; however, most define CPP as intermittent or constant, localised to the lower abdomen or pelvis and of at least 6 months duration^3^. CPP is the single most common presentation to gynaecology services, implicated in 20% of outpatient gynaecology appointments^1^ and > 40% of laparoscopies^4^. There are numerous causes of CPP, including endometriosis, and many cases of CPP are considered multifactorial^5^.

Endometriosis is estimated to affect 1 in 10 Australian women of reproductive age and is calculated to cost >$6billion/annum in direct medical and surgical costs for women >18 years of age alone^5^.The gold standard for endometriosis diagnosis requires laparoscopy and histopathology^3,6^. For many women, the delay between first symptoms and diagnosis can be >8 years^6^. As a result, there is keen interest to identify clinical features that may predict the finding of endometriosis and mitigate the delay in initiating active treatment^7^.

Symptoms attributed to endometriosis usually relate to infertility and pain, although there is evidence that endometriosis can be asymptomatic^6^. The reported pain with endometriosis may consist of dysmenorrhoea, dyspareunia and non-cyclic pelvic or abdominal pain^8^. A recent review of studies exploring potential symptoms of endometriosis failed to find any symptom, symptom cluster or algorithm that was predictive^9^. For women with CPP undergoing laparoscopy, approximately 30% are diagnosed with endometriosis, however 30-50% will have no pathology identified^3^. Additionally, there is conflicting evidence regarding any correlation between symptom intensity and endometriosis stage^9,10^. Furthermore, the significance of mild endometriosis and the utility of surgical removal for pain management is questionable^11^. This also reflects the increasing recognition of the role of central pain sensitisation in pain responses^12^ and helps to explain the co-existence of other pain syndromes in both people with CPP and those with endometriosis. In addition to these intrinsic patient factors, the impact of the clinician on the decision making processes regarding treatment, has received minimal academic consideration^13^.

This study aims to explore the characteristics of women referred with CPP and to identify any correlation between their pain symptoms, the undertaking of surgery and the diagnosis and stage of endometriosis.

## Methods

### Participants and setting

This prospective observational study is part of a larger cohort study, the Persistent Pelvic Pain(PPP) project (ANZCTR:ACTRN12616000150448), which has been conducted in a public women’s hospital in Melbourne, Australia. The trial protocol has been published^14^. Participants were enrolled following referral to outpatient clinics if their referral indicated any pain symptoms (dysmenorrhoea, non-cyclic pelvic pain, dysuria, dyspareunia and/or dyschezia) and they were aged between 18-50 years. Exclusion criteria were:

a. Referral documented subfertility or pregnancy planning
b. Recent sonographic evidence of ovarian cyst(>5cm)
c. Prior hysterectomy

### Study design

Ethics approval (HREC R14-31) was granted. Informed consent was obtained from all participants. Women were randomised to one of two gynaecology clinics for routine ongoing gynaecology care. Participants were followed for three-years with surveys administered 6-monthly and their medical records were reviewed to obtain relevant examination, operative and investigation findings (Figure 1).

**Figure 1:**
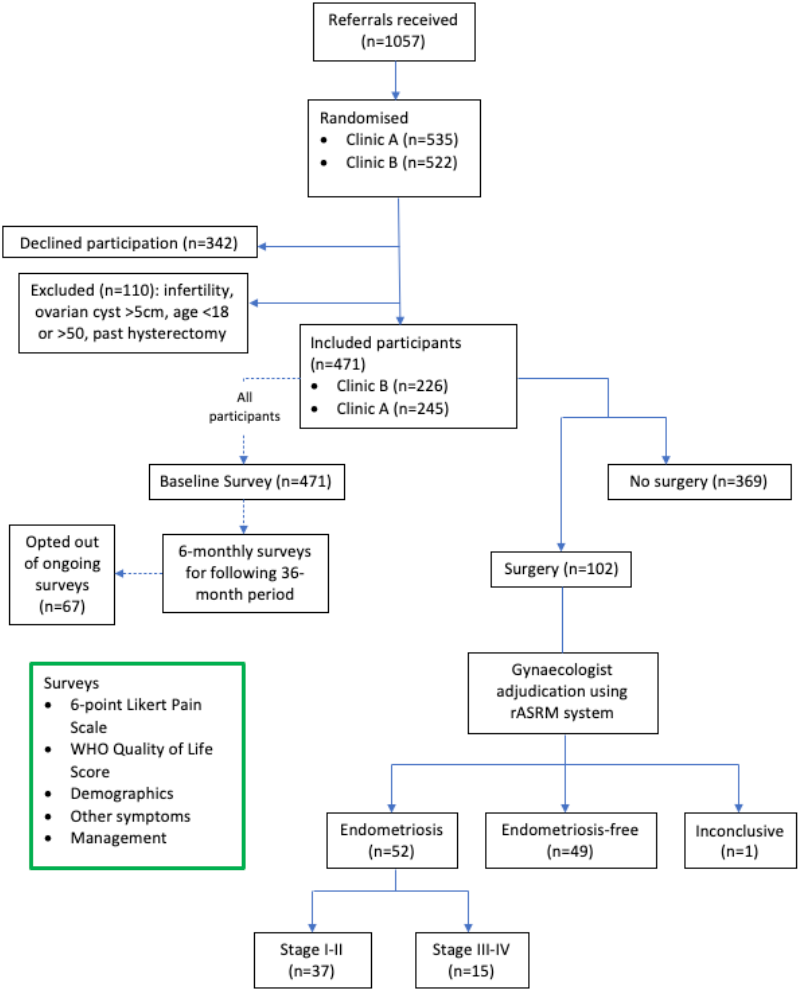
Flow chart of study recruitment, survey follow up and surgery.

Participants completed baseline surveys exploring demographics, past medical history, World Health Organization Quality of Life Bref score (WHO QoL-Bref) and self-reported pain scores rated on a Likert scale. Gynaecologists treating study participants were blinded to questionnaire responses. The final, 36-month survey was accepted within a 6-month window of this date.

Operation reports identifying endometriosis were retrospectively staged using revised American Society Reproductive Medicine(rASRM) by a single gynaecologist blinded to other information.

### Outcome measures

This study examined characteristics and potential factors that might influence three questions:

1. Who has surgery?
2. Who is diagnosed with endometriosis?
3. What influences endometriosis severity?

Baseline and nearest pre-operative surveys were used to explore potential predictive factors for these questions. Age, age of symptom onset, menarche, gravidity, parity, QoL, self-rated pain scores and family history of period pain, heavy periods or endometriosis were analysed in this study.

#### Pain Score

Patients rated their average pain scores over the last three months at each survey, for dysmenorrhoea, non-cyclic pelvic pain, dysuria, dyspareunia, dyschezia and satisfaction with overall pain control on a 6-point Likert scale (0-5). A score of 0 was no pain at all and 5, the worst pain imaginable. For the satisfaction scale, 0 was determined to be completely unsatisfied, whilst 5 was completely satisfied. The satisfaction scale was recoded to represent dissatisfaction and contributed to the derivation of an average total pain score.

#### Gynaecology unit

Two general gynaecology units were involved: one with additional advanced endoscopic skills (clinic A) and the other with additional hormonal and pain management skills (clinic B).

### Data analysis and statistics

Baseline survey responses completed by all participants were used to determine predictors of surgical intervention. In the surgical group, for analysis of the predictors of endometriosis and endometriosis severity, nearest pre-operative survey responses were analysed for QoL and pain scores.

Statistical analysis was performed using STATA/IC 16.1, StataCorp LLC. Parametric and non-parametric tests were performed where appropriate. To analyse potential predictors of surgery, endometriosis and endometriosis severity, a multivariate logistic regression model was designed. Penalised Maximum Likelihood Estimation (PMLE) 15 or exact logistic regression models were used where the number of observations was less than 100. All independent variables underwent univariate logistic regression against the binary outcome variable, with a cut-off score of P< 0.10 for progression to the regression model^16^. For univariate regressions with less than 200 observations, exact logistic regression was used. Variables deemed to be clinically or statistically collinear (variance inflation factor (VIF) > 10^17^) were identified and removed where necessary. Likert subscales were treated as continuous independent variables for the purpose of analysis, providing they encompassed at least 5 values and approximated linear scales. Missing data was explored and 10% was set as the cut-off for progression to multiple imputation^18^To account for missing data in individual responses to pain parameters contributing to a total pain score, an overall median pain score was used. Results of the multivariate regression are presented as odds ratios (OR) with 95% confidence intervals (CI). Statistical significance was set at P<0.05.

## Results

### Baseline characteristics

Of the1057 referrals for pelvic pain, 471 met the inclusion criteria and consented to participation. The baseline demographics of the study cohort are detailed in Table 1.

**Table 1:** Baseline characteristics of women recruited to the PPP study.

| Parameter | Study cohort | Clinic A | Clinic B | P-value |
| --- | --- | --- | --- | --- |
| **Number Participants** | 471 | 245 | 226 |  |
| **No surgery** | 369 | 173 | 196 |  |
| **Surgery** | 102 | 72 | 30 | <0.001* |
| **No endometriosis** | 49 | 35 | 14 |  |
| **Endometriosis** | 52 | 37 | 15 | 0.976 |
| **Endometriosis: stage I-II** | 37 | 25 | 12 |  |
| **Endometriosis: stage III-IV** | 15 | 12 | 3 | 0.370 |
| **Age at referral (years)** | 30 [24, 38] | 32 [26, 40] | 28 [23, 36] | <0.001* |
| **Age of menarche** | 12.5 [11.5, 14] | 13 [12, 14] | 12 [11, 14] | 0.317 |
| **Family history of period pain, n(%)** | 274 (63.13%) | 139 (61.8%) | 135 (62.5%) | 0.876 |
| **Family history of heavy periods, n(%)** | 240 (54.30%) | 115 (51.3%) | 125 (57.3%) | 0.206 |
| **Family history of endometriosis, n(%)** | 148 (33.48%) | 78 (35.0%) | 70 (32.0%) | 0.502 |
| **Gravidity** | 1 [0, 3] | 1 [0, 3] | 0 [0, 3] | 0.143 |
| **Parity** | 0 [0, 2] | 0 [0, 2] | 0 [0, 2] | 0.087 |
| **Total WHO-QoLBref** | 89 [77, 100] | 91 [77, 100] | 87 [76, 98] | 0.112 |
| **Dysmenorrhoea** | 4 [3, 5] | 4 [3, 5] | 4 [3, 4] | 0.927 |
| **Non-cyclic pelvic pain** | 4 [2, 4] | 4 [2, 4] | 3 [2, 4] | 0.757 |
| **Dysuria** | 0 [0, 2] | 1 [0, 2] | 0 [0, 2] | 0.461 |
| **Dyspareunia** | 3 [1, 4] | 3 [1, 4] | 3 [1, 4] | 0.749 |
| **Dyschezia** | 2 [0, 3] | 2 [0, 3] | 2 [0, 3] | 0.725 |
| **Dissatisfaction with pain management** | 3 [2, 4] | 3 [2, 4] | 3 [2, 4] | 0.821 |
| **Overall median pain score** | 3 [1.5, 3.5] | 2.5 [2, 3.5] | 2.5 [1.5, 3.5] | 0.923 |

PPP = Persistent pelvic pain, WHO-QoLBref = World Health Organization Quality of Life Bref
Values are given as medians [25<sup>th</sup> and 75<sup>th</sup> percentiles] and number (% of responses). Concerning the entire study cohort, all responses are derived from baseline surveys. Concerning the two clinics, pain, QoL and pain catastrophising scores are derived from nearest pre-operative surveys. P-values are derived from comparison of Clinic A and B patients.
\* Denotes statistical significance.

Of those recruited, median age was 30years and median age of symptom onset was 20-29years.

Median age of menarche was 12.5years and many women reported a family history of period pain, heavy periods or endometriosis (63%, 54% and 33% respectively). The overall median pain score reported was 3 on the Likert scale. Women randomised to clinic A and B differed only by age at referral (32 and 28 years respectively, P<0.001) and by number of surgical procedures (clinic A > clinic B (P<0.001)).

During the 36-month follow-up, 102 patients had either a laparoscopy(n=100) or laparotomy(n=2). On review of operation reports, 52(51%) had endometriosis identified and 49 were negative for endometriosis, whilst one report was inconclusive. Using the rASRM, 37 patients had stage I-II endometriosis whilst 15 had stage III-IV endometriosis.

### Predictors for undertaking surgery

The variables identified as potential predictors of undergoing surgery were gynaecology unit, parity, dysmenorrhoea, non-cyclic pelvic pain, dysuria, dyschezia and overall median pain score. Considering potential collinearity and model simplicity, the overall median pain score was used in lieu of the individual domain pain scores. This study demonstrated that there was a 65% reduction in the odds of undergoing surgery for women allocated to Clinic B when compared to women in Clinic A (OR 0.342; 95%CI: 0.209-0.561, P<0.001). There was a 24% decrease in likelihood of having surgery for every child a woman had (OR 0.767; 95%CI: 0.620-0.949, P=0.015). Additionally, a 30% increase in the odds of undergoing surgery was determined for every one-point increase in median pain score (OR 1.303; 95%CI: 1.079-1.573, P=0.006). These results are provided in Table 2a.

**Table 2a:** Baseline predictors for surgery (laparoscopy or laparotomy) (n=439)

| Predictors of Surgery | Odds ratio | 95% Confidence Interval | P- Value |
| --- | --- | --- | --- |
| **Clinic (A = 0, B = 1)** | 0.342 | [0.209-0.561] | <0.001 |
| **Parity** | 0.767 | [0.620-0.949] | 0.015 |
| **Overall median pain score** | 1.303 | [1.079-1.572] | 0.006 |

Multivariate logistic regression. Clinic allocation was coded as 0= Clinic A, 1= Clinic B.

### Predictors of finding endometriosis at surgery

Referral age, gravidity, parity and dissatisfaction with pain management were identified as potential predictors of endometriosis diagnosis in the surgical cohort of participants (see Appendix A). As parity is a subset score of gravidity, these variables were considered clinically collinear and gravidity was excluded in multivariate regression. The allocated unit did not predict the discovery of endometriosis at surgery (OR 1.012; 95%CI: 0.392-2.634, P=1.000).

As shown in Table 2b, within the PPP cohort of women who had surgery, no statistically significant predictors of finding endometriosis were identified(P=0.036).

**Table 2b:** Preoperative predictors of endometriosis (n=97)

| Predictors of Endometriosis | Odds ratio | 95% Confidence Interval | P- Value |
| --- | --- | --- | --- |
| Age | 0.972 | [0.917-1.032] | 0.353 |
| Parity | 0.682 | [0.396-1.178] | 0.169 |
| Dissatisfaction with pain management | 1.266 | [0.907-1.767] | 0.166 |

Penalised Maximum Likelihood Estimation

### Predictors of endometriosis severity at surgery

Appendix A demonstrates that age, family history of heavy periods, family history of endometriosis and total WHO-QoL score were potential predictors for endometriosis severity. Due to its low OR effect size (OR 1.049; 95%CI: 1.005-1.101) and strong clinical and statistical collinearity with pain score, total WHO-QoL score was excluded from the multivariate regression model.

Age was a statistically significant predictor of endometriosis severity(P=0.003), indicating a 15% increase in the odds of Stage III-IV over Stage I-II endometriosis with each one-year increase in age (see Table 2c). No other predictors of endometriosis severity were identified.

**Table 2c:**
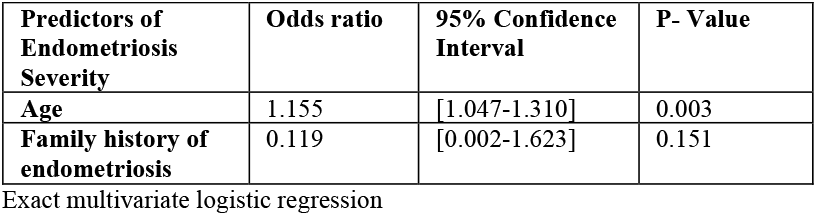
Preoperative predictors of endometriosis severity (n=47)

| Predictors of Endometriosis Severity | Odds ratio | 95% Confidence Interval | P- Value |
| --- | --- | --- | --- |
| Age | 1.155 | [1.047-1.310] | 0.003 |
| Family history of endometriosis | 0.119 | [0.002-1.623] | 0.151 |

Exact multivariate logistic regression

## Discussion

The purpose of this study was to explore the predictors for undergoing surgery, for identifying endometriosis and endometriosis severity in a cohort of women with CPP. As a cohort observational study, actual clinical care was not influenced by the study. Thus, the results are “real-world” outcomes, avoiding the problems arising from randomised controlled studies where the participant cohort may be biased by their interest in participating in research and intensely followed up, often with regular contact from the research team^19^.

The primary findings of this study include:

1. Women attending a gynaecology unit with specific additional endoscopic skills are more likely to undergo surgery than women with similar symptoms who attend a unit with additional skills in medical management.
2. Women with CPP who had fewer children and reported more pain were more likely to undergo surgery.
3. 51% of women who underwent laparoscopy were found to have endometriosis.
4. No parameters assessed in this study were predictive of endometriosis diagnosis in women who underwent surgery. Notably, pain was ***not*** determined to be a likely predictor of endometriosis diagnosis.
5. In our cohort, older age increased the likelihood of being diagnosed with more severe endometriosis. No other significant predictors were identified. Notably, pain was ***not*** determined to be a likely predictor of endometriosis severity.

The impact of the gynaecologist in the routine clinical discussions with the patient has not previously been identified, although suspected^13^. The real-world approach of this observational study of a randomised patient cohort, allowed clinical care provided under two different styles of practice to be assessed. This could not easily be undertaken as a prospective randomised controlled trial, which would need to incorporate randomisation of clinician behaviour and the study findings would be impacted by the artificial personalisation of follow-up processes required of such studies^19^. It was clear from this study that the clinic to which patients were randomised significantly impacted on their likelihood of surgery, favouring operation if randomised to the unit with specialist endoscopic surgeons. However, the unit allocation did not serve as a predictor of finding endometriosis or of having more severe endometriosis at surgery. Whilst both pain and unit allocation were predictors of undergoing surgery, these findings appear to be independent of one another as there was no difference in pain scores across units. It seems these findings indicate that the specialisation and training of the gynaecologist has a significant impact on the decision to operate.

A previous PPP publication reporting on the characteristics of women who underwent laparoscopy in the first 211 participants, demonstrated that only severe dysmenorrhoea predicted surgery^20^. Analysis of data by gynaecology unit was not performed to avoid potentially influencing clinician behaviour in the remainder of the study.

Our study indicated that women who had fewer children were more likely to have surgery. This is difficult to explain, as subfertility was an exclusion criterion for the study and age was not a significant predictor of likelihood of surgery.

The endometriosis rate in our study of 51% is consistent with rates reported in pain populations^21^. Higher-powered studies and reviews which question study definitions of CPP, report a lower rate (30%) of endometriosis amongst women with CPP^22^. Furthermore, a study exploring the prevalence of endometriosis and pre-surgical symptoms indicated that the prevalence of endometriosis in asymptomatic women (15.2%) was similar to the prevalence in a CPP population (21.4%)^21^.

The lack of pain symptoms that predict visualised endometriosis in this study is important, supporting a recent review^9^.This highlights the need to review current guidelines that advise that pain intensity is an indication for referral for laparoscopy.

In addition to the lack of correlation between pain severity and endometriosis diagnosis, our study also failed to show a correlation between pain intensity and endometriosis stage. The literature has conflicting findings relating to symptom severity and endometriosis severity^10,23,24,25^.

There are a number of limitations affecting interpretation of this study. Missing responses to a number of variables was noted, although no missing responses exceeded 10% and given the cohort size, this was deemed acceptable for analysis. Individual missing variables contributing to overall category scores was a problem. For pain scores, a median score across the six survey domains of pain was taken for each individual participant to account for missing responses to some of these variables. This was more complicated when exploring the WHO-Qol scores, where omitted answers to all questions in a section of the survey impacts the numerical total score which relates directly to the interpretation of the validated scale. This could not be directly adjusted for and as such may have impacted on the predictability of these outcomes for surgery, endometriosis and endometriosis severity.

Whilst the whole PPP study cohort is quite large, the proportion who had surgery and had endometriosis was substantially smaller. To account for this, alternative statistical methods proven to be better for smaller case numbers were used, however, some caution with interpretation of these results is still required.

This study reiterates the need to expand our understanding of pelvic pain and to continue to question the association between pain and endometriosis. Our findings support existing literature that pain intensity is a predictor of the likelihood of undergoing surgery. However, in contradiction to this finding, pain intensity was not a predictor of the presence of endometriosis at surgery and did not correlate to the severity of endometriosis found. These findings need to be considered in the development of future guidelines.

Our study findings re-emphasise the need to address pain concerns directly, with caution in approaching laparoscopy as means to diagnosis and treatment. Almost half of the participants showed no diagnosable endometriosis pathology, despite clinically indistinguishable pain symptoms. Laparoscopy comes at a cost and most certainly is not without risk. These findings, along with the evidence indicating no direct association between endometriosis and pain, should be motivation to continue research that seeks to address pelvic pain.

## Data Availability

The datasets generated for this study are available from the corresponding author on reasonable request.

## Acknowledgements

We acknowledge the financial support from the Norman Beischer Medical Research Foundation, The Pelvic Pain Foundation Australia (https://www.pelvicpain.org.au) and the Mercy Hospital Victoria Limited Small Research Grants.

None of the authors hold any conflict of interest.

## Appendix A

Univariate logistic regression analyses of potential predictors for surgery, endometriosis and endometriosis severity

| Variable | No surgery vs Surgery |  |  | No Endometriosis vs Endometriosis |  |  | Endometriosis: Stage I-II vs Stage III-IV |  |  |
| --- | --- | --- | --- | --- | --- | --- | --- | --- | --- |
|  | *Odds ratio [95% CI]* | *P-value* | *No. obs* | *Odds ratio [95% CI]* | *P-value* | *No. obs* | *Odds ratio [95% CI]* | *P-value* | *No. obs* |
| **Clinic (A =0, B =1)** | 0.368 [0.229-0.590] | 0.000 | 471 | 1.013 [0.392-2.634] | 1.000 | 101 | 0.527 [0.080-2.503] | 0.588 | 52 |
| **Age at referral** | 0.995 [0.970-1.021] | 0.711 | 470 | 0.946 [0.900-0.993] | 0.025 | 101 | 1.205 [1.093-1.357] | 0.000 | 52 |
| **Age of menarche** | 0.967 [0.863-1.084] | 0.565 | 463 | 0.960 [0.752-1.221] | 0.759 | 98 | 1.250 [0.857-1.857] | 0.256 | 50 |
| **FHx period pain** | 1.420 [0.876-2.033] | 0.155 | 441 | 0.963 [0.365-2.530] | 1.000 | 94 | 0.316 [0.068-1.404] | 0.149 | 48 |
| **FHx heavy periods** | 1.272 [0.803-2.015] | 0.305 | 442 | 1.525 [0.618-3.812] | 0.424 | 94 | 0.179 [0.035-0.792] | 0.020 | 48 |
| **FHx endometriosis** | 1.234 [0.768-1.983] | 0.386 | 442 | 0.988 [0.393-2.490] | 1.000 | 93 | 0.080 [0.002-0.648] | 0.010 | 48 |
| **Gravidity** | 0.933 [0.816-1.067] | 0.311 | 444 | 0.672 [0.486-0.901] | 0.006 | 98 | 1.302 [0.748-2.251] | 0.373 | 50 |
| **Parity** | 0.802 [0.654-0.985] | 0.035 | 443 | 0.543 [0.323-0.870] | 0.009 | 97 | 1.798 [0.795-4.107] | 0.170 | 49 |
| **Total WHO QoL score** | 0.993 [0.979-1.006] | 0.287 | 462 | 0.997 [0.973-1.021] | 0.811 | 100 | 1.049 [1.005-1.101] | 0.026 | 52 |
| **Dysmenorrhoea** | 1.207 [1.001-1.457] | 0.049 | 455 | 0.987 [0.996-1.348] | 0.996 | 98 | 0.750 [0.470-1.175] | 0.220 | 52 |
| **Non-cyclic pelvic pain** | 1.189 [1.024-1.381] | 0.023 | 467 | 0.799 [0.590-1.066] | 0.134 | 101 | 0.934 [0.611-1.441] | 0.810 | 52 |
| **Dysuria** | 1.142 [0.990-1.316] | 0.068 | 466 | 0.990 [0.760-1.290] | 0.990 | 101 | 0.967 [0.618-1.482] | 0.965 | 52 |
| **Dyspareunia** | 1.078 [0.947-1.228] | 0.252 | 433 | 0.986 [0.777-1.250] | 0.952 | 97 | 0.901 [0.620-1.304] | 0.624 | 50 |
| **Dyschezia** | 1.139 [1.000-1.298] | 0.050 | 465 | 1.052 [0.828-1.339] | 0.714 | 101 | 1.037 [0.716-1.512] | 0.914 | 52 |
| **Dissatisfaction with pain** | 1.124 [0.945-1.338] | 0.186 | 467 | 1.327 [0.964-1.848] | 0.085 | 101 | 0.902 [0.533-1.534] | 0.770 | 52 |
| **Overall median pain score** | 1.300 [1.088-1.554] | 0.004 | 467 | 0.847 [0.604-1.178] | 0.337 | 101 | 0.860 [0.517-1.421] | 0.584 | 52 |
| **Total pain catastrophising score** | 1.011 [0.994-1.029] | 0.211 | 451 | 1.006 [0.976-1.036] | 0.713 | 99 | 0.954 [0.906-1.000] | 0.049 | 51 |

Survey responses to the baseline survey are used for demographic questions and for predictors of undergoing surgery. World Health Organization Quality of Life Bref score (WHO-QoLBref), pain scores and pain catastrophising scores predicting endometriosis and endometriosis severity are taken from the nearest pre-operative survey. FHx= family history. Clinic allocation was coded as 0= Clinic A, 1= Clinic B.

